# Inpatient COVID-19 Mortality Rates: What are the predictors?

**DOI:** 10.1101/2022.01.07.22268906

**Authors:** Mona Al-Amin, Md Nazmul Islam, Kate Li, Natalie E. Sheils, John Buresh

## Abstract

**Objective:** This study aims to investigate the relationship between registered nurses and hospital-based medical specialties staffing levels with inpatient COVID-19 mortality rates.

**Methods:** We rely on data from AHA Annual Survey Database, Area Health Resource File, and UnitedHealth Group Clinical Discovery Database. We use linear regression to analyze the association between hospital staffing levels and bed capacity with inpatient COVID-19 mortality rates from March 1, 2020, through December 31, 2020.

**Results:** Higher staffing levels of registered nurses, hospitalists, and emergency medicine physicians were associated with lower COVID-19 mortality rates. Moreover, a higher number of ICU and skilled nursing beds were associated with better patient outcomes. Hospitals located in urban counties with high infection rates had the worst patient mortality rates.

**Conclusion:** Higher staffing levels are associated with lower inpatient mortality rates for COVID-19 patients. A future assessment is needed to establish benchmarks on the minimum staffing levels for nursing and hospital-based medical specialties during pandemics.

## Introduction

COVID-19 is a serious infection that spreads rapidly within a population and has placed tremendous strain on hospitals worldwide. As of August 2021, around 650,000 people infected with COVID-19 have lost their lives in the U.S. alone. While 80% of individuals infected with the virus exhibit mild symptoms, around 20% produce a “hyperinflammatory response” and suffer from severe respiratory distress.^1,2^ Estimates of case fatality vary between countries and even within single countries. For instance, Ondor et al. (2020) estimated that the case fatality rate in Italy was 7.20%.^3^ While Verity et al. (2020) estimated an overall fatality rate of 1.38% in China. ^4^ He et al., based on a meta-analysis and sensitivity analysis, estimate an overall case fatality rate of 2.72%.^5^ Patient characteristics explain some of this variability. Price-Haywood et al. found that patients who were Black, older, on Medicare or Medicaid, obese, or resided in lower-income neighborhoods were more likely to be admitted to a hospital.^6^ Among these patient characteristics, only older age, was associated with higher in-hospital mortality.

While patient characteristics help explain some of the variability, hospital characteristics, such as capacity, and county-level variables, such as infection rates, are likely to influence COVID-19 mortality rates. Strained capacity, specifically, limits hospitals in their response to the “pandemic-associated surge” and contributes to mortality from COVID-19.^7^ Data from the epicenter of the pandemic provides evidence that there is variation in mortality rates even within the same region.^8^ According to Ji et al., this variation can be explained by the impact the availability of healthcare resources has on COVID-19 mortality rates.^8^

Limited research examined hospital-level predictors of COVID-19 mortality. As Asch et al. argue; “hospital-level mortality may depend not just on patient risk factors, but also on the hospital where patients are admitted.”^9^ When pandemics occur, hospitals face a significant increase in demand in a short period of time which has an impact on patient outcomes. Demand surge is defined as the rapid response to “meet increased demand in medical care.”^10^ During public health emergencies, such as pandemics, capacity, specifically beds, staff, and equipment experience “severe strain resulting from patient surges.”^11^ There are four key elements, referred to as the 4S, to surge capacity which include staff, stuff (beds and equipment), structure (space), and management systems.^12,13^ The availability of hospital beds and equipment in addition to sufficient staffing levels are essential elements of any response to increased demand in healthcare.^14^ Staffing capacity impacts the workload experienced by clinical staff which is associated with patient outcomes.^15^ Additionally, hospital staffing levels impact patient outcomes indirectly via the significant impact on healthcare workers’ psychological well-being. As Niels et al. argue, the psychological burden that healthcare workers endure during a pandemic should be considered when making staffing decisions.^16^

Hospital capacity is likely to influence patient outcomes during a pandemic by causing delays in admissions, diagnosis, treatment, and/or transfer to the intensive care unit (ICU). The objective of this paper is to study the association between hospital capacity and county level variables with COVID-19 patient outcomes at the hospital level. We focus on the staff dimension of surge capacity by investigating the relationship between baseline staffing levels of registered nurses (RNs), hospitalists, emergency medicine physicians, and intensivists (staff), with hospital-level COVID-19 mortality rates. This paper builds on and expands the model used by Asch et al.^9^ who only examined a limited set of hospital level variables over a shorter period of time.

## Methods

### Data Sources

We used hospital□level data from the 2019 American Hospital Association (AHA) Annual Survey Database, de-identified patient level data from the UnitedHealth Group Clinical Discovery Database, and county-level data from the 2019-2020 Area Health Resource File (AHRF). We also obtained county-level cumulative COVID-19 case rates for March 1 to December 31, 2020 from *The New York Times* database.

We used de-identified administrative claims from the UnitedHealth Group Clinical Discovery Database for both Medicare and commercial fully insured populations (including patients with administrative services (ASO) only) accompanied by a daily record of hospital admissions due to primary or secondary diagnosis of COVID-19 along with the daily disposition status (admitted, discharged, transferred, or expired) available until December 31, 2020. Data for commercial populations included both fully-insured members and, where available in the research database, administrative services (ASO) populations. The three datasets were merged at the hospital level to identify predictors of COVID-19 mortality rates. We limit our study to not□for□profit, for□profit, and nonfederal public general hospitals and hospitals with more than or equal to 10 patients from our database admitted with COVID-19 diagnosis during the study period. We analyzed data in two phases: while the patient-level analysis (phase-1) was based on 1,397 hospitals exploiting 95,919 patients, the hospital-level analysis (phase-2) consisted of 1,370 hospitals with no missing hospital- and county-level data; for details, please refer to Figure SM4. This study was reviewed and deemed exempt by the institutional review board of UnitedHealth Group.

### Dependent Variable

The primary outcome was hospital specific risk standardized event rates (RSERs) signaling estimated inpatient mortality or referral to hospice within 30-days of admission; the rates in probability scale were adjusted for patient-level characteristics (e.g., demographics, Elixhauser comorbidities,^17^ community or nursing facility admission), hospital size with respect to average patient volume seen per day based on 2018-19 census data (AHA Annual Survey Database) and discretized time points featuring different pandemic phases with respect to the differences between patients’ first hospital admission and March 1, 2020.

To create the study population, we included all Medicare Advantage and commercially insured hospitalized patients, including data for ASO plan members where available in the database. Next, we restricted the sample to patients continuously enrolled for at least six months to capture better the claims-based historical comorbidities. We further confined the sample to patients who are older than eighteen and excluded any cases with missing patient-level risk factors. To accommodate two pandemic surges that happened in 2020, we excluded patients admitted before March 1, 2020 and after December 31, 2020. We included hospitals only with medical provider numbers matched to the AHA database. Patients with readmission or a history of hospital transfer within 30-days of the initial admission are precluded to evade any potential misattribution of hospital-level outcomes; a sensitivity analysis relaxing this criterion is also performed. To improve the robustness of hospital-specific parameter estimates, we included sites with at least ten patients who are present in our database; see Figure SM5 for details.

### Predictors

#### Hospital Level Variables

The primary objective of this paper is to study the association between hospital capacity and county level variables with COVID-19 mortality rates at the level of the hospital. We focus on the number of beds and staff dimensions of hospital surge capacity. For bed capacity, we include the number of hospital beds, intensive-care unit (ICU) beds, and skilled nursing care beds. For staffing capacity, we capture baseline staffing levels of RNs, hospitalists, intensivists, and emergency medicine physicians from 2019. Also at the hospital level, we control for ownership (not-for-profit, for-profit, and local public hospitals), whether a hospital has one or more Accreditation Council for Graduate Medical Education (ACGME) accredited programs, whether a hospital belongs to a system, and baseline utilization (calculated as the number of inpatient days divided by 365 multiplied by the number of hospital beds in 2019).

#### County Level Variables

For county level variables, we control for location (urban vs. rural), percentage of persons in poverty, percentage of Black/African American, percentage of Hispanic/Latino, and the cumulative COVID case rate per 10,000 residents during March-December 2020.

### Statistical Analysis

Descriptive statistics of the patient-level and hospital-level variables are stipulated in Table SM2 and Table 1, respectively. To select a set of influential variables and avoid over-fitting, a variable screening step based on univariate generalized linear model is performed with respect to each covariate of interest.^18^ Hierarchical model is fitted to estimate the odds of mortality or referral to hospice adjusting for the selected covariates including Elixhauser based comorbidities, demographic variables (age and gender), status of transfer from nursing facility admission, the number of days between each hospital admission and March 1, 2020, and the volume of average patient admissions in 2018-19; the last variable is to adjust for the association between hospital volume and mortality rates.Variation among patients is accounted for through hospital specific random effects which are estimated via restricted maximum likelihood (REML). We report the adjusted odds ratios of each risk factors with 95% confidence intervals along with p-values based on Wald test statistics. Each hospital’s RSERs were calculated via recycled prediction where the idea is to take average over all patients’ predicted probabilities of experiencing events had each of them hypothetically been treated at each hospital; for details we refer to Asch, et al.^9^, George, et al.^18^, Silber, et al.^19-21^ Second model is fitted by excluding patients with readmission or transfer to facilities that are only short-term, long-term, and critical access care; Kendall rank correlation coefficient was used to compare the differences in ranking of hospitals with respect to the corresponding RSERs. Technical details are provided in the supplementary material.

**Table 1:**
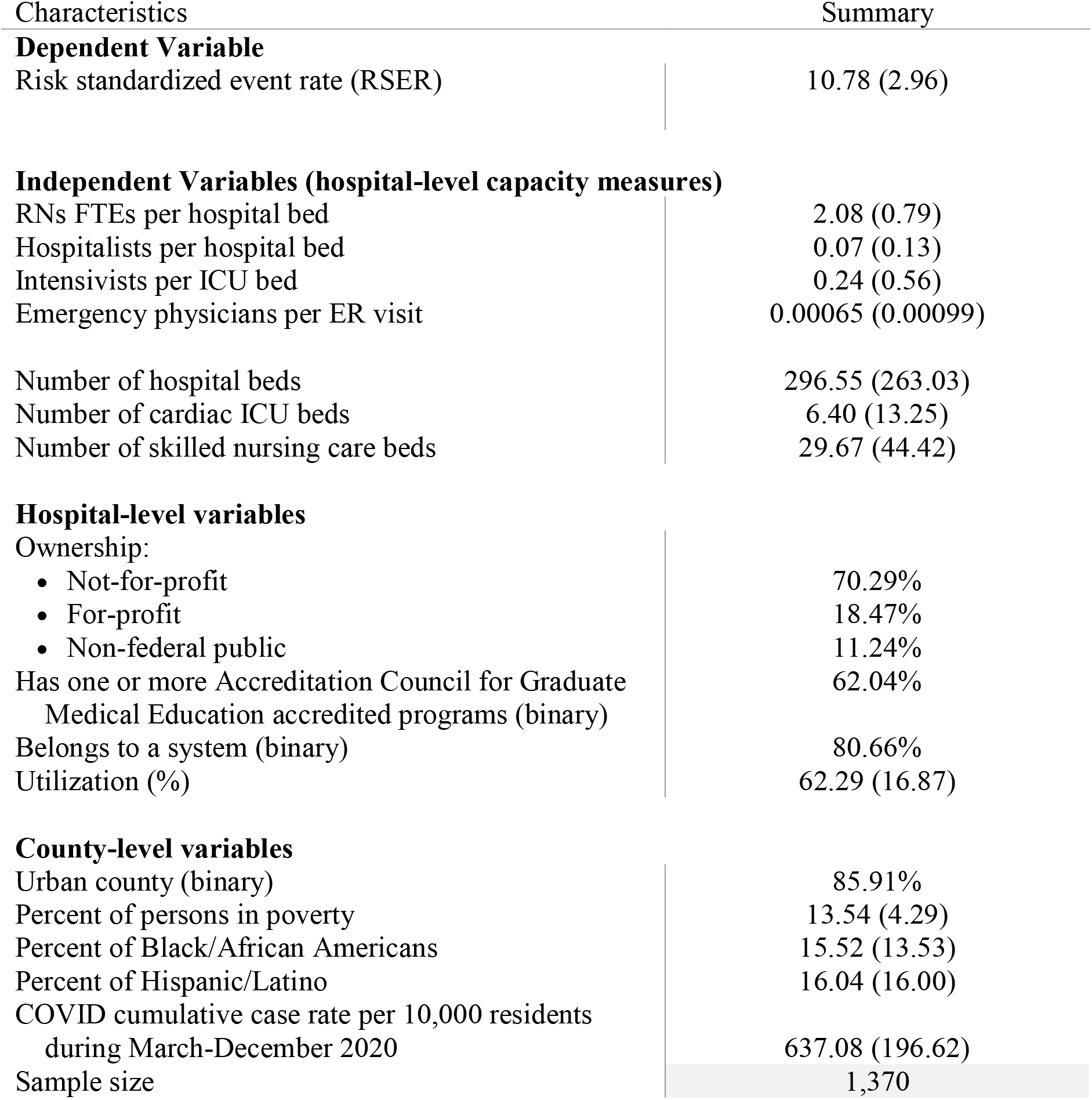
Baseline characteristics for 1,370 hospitals. Reported are the mean and standard deviation for numeric variables, and count and percentage (%) for categorical variables.

| Characteristics | Summary |
| --- | --- |
| **Dependent Variable** |  |
| Risk standardized event rate (RSER) | 10.78 (2.96) |
| **Independent Variables (hospital-level capacity measures)** |  |
| RNs FTEs per hospital bed | 2.08 (0.79) |
| Hospitalists per hospital bed | 0.07 (0.13) |
| Intensivists per ICU bed | 0.24 (0.56) |
| Emergency physicians per ER visit | 0.00065 (0.00099) |
| Number of hospital beds | 296.55 (263.03) |
| Number of cardiac ICU beds | 6.40 (13.25) |
| Number of skilled nursing care beds | 29.67 (44.42) |
| **Hospital-level variables** |  |
| Ownership: |  |
| • Not-for-profit | 70.29% |
| • For-profit | 18.47% |
| • Non-federal public | 11.24% |
| Has one or more Accreditation Council for Graduate Medical Education accredited programs (binary) | 62.04% |
| Belongs to a system (binary) | 80.66% |
| Utilization (%) | 62.29 (16.87) |
| **County-level variables** |  |
| Urban county (binary) | 85.91% |
| Percent of persons in poverty | 13.54 (4.29) |
| Percent of Black/African Americans | 15.52 (13.53) |
| Percent of Hispanic/Latino | 16.04 (16.00) |
| COVID cumulative case rate per 10,000 residents during March-December 2020 | 637.08 (196.62) |
| Sample size | 1,370 |

We treat the estimated RSER as the dependent variable in the subsequent linear regression model which is used to investigate the association between hospital capacity and county level variables with COVID-19 mortality rates. State level fixed effects are added in the model to account for any potential unobserved heterogeneity. All statistical tests were two-sided with 5% significance level and reported p-values were not adjusted for multiplicity. All analyses were conducted using R version 3.6.3.

## Results

Adjusted ORs associated with the risk factors that were used in calculating RSERs are illustrated in Figure SM1. Figure SM2 shows the estimated RSERs in an ascending order with the corresponding interquartile range (IQR); here smaller RSER indicates lower rate of experiencing events at hospital-level.

The results of the hospital level linear regression model are shown in Table 2. Based on the diagnostic plots of the regression model, the model assumptions were largely met. All variance inflation factors (VIFs) were less than 3.30, which suggested that multicollinearity was not a concern. Among the four staffing variables, RNs (estimate, -0.32; 95% CI, -0.50 – -0.14), hospitalists (estimate, -0.16; 95% CI, -0.33 – 0.00), and emergency medicine physicians’ levels (estimate, -0.14; 95% CI, -0.30 – 0.02) are negatively correlated with RSER indicating that higher staffing level of these three types are associated with lower COVID-19 mortality rate. On the contrary, increased intensivists staffing levels are associated with higher mortality rate. As for bed capacity, higher number of hospital beds (estimate, 0.49; 95% CI, 0.26–0.72) is associated with higher mortality rate, while increases in the number of cardiac ICU beds (estimate, -0.16; 95% CI -0.36–0.03) and the number of skilled nursing care beds (estimate, - 0.22; 95% CI, -0.39 – -0.06) are associated with lower mortality rate.

**Table 2:**
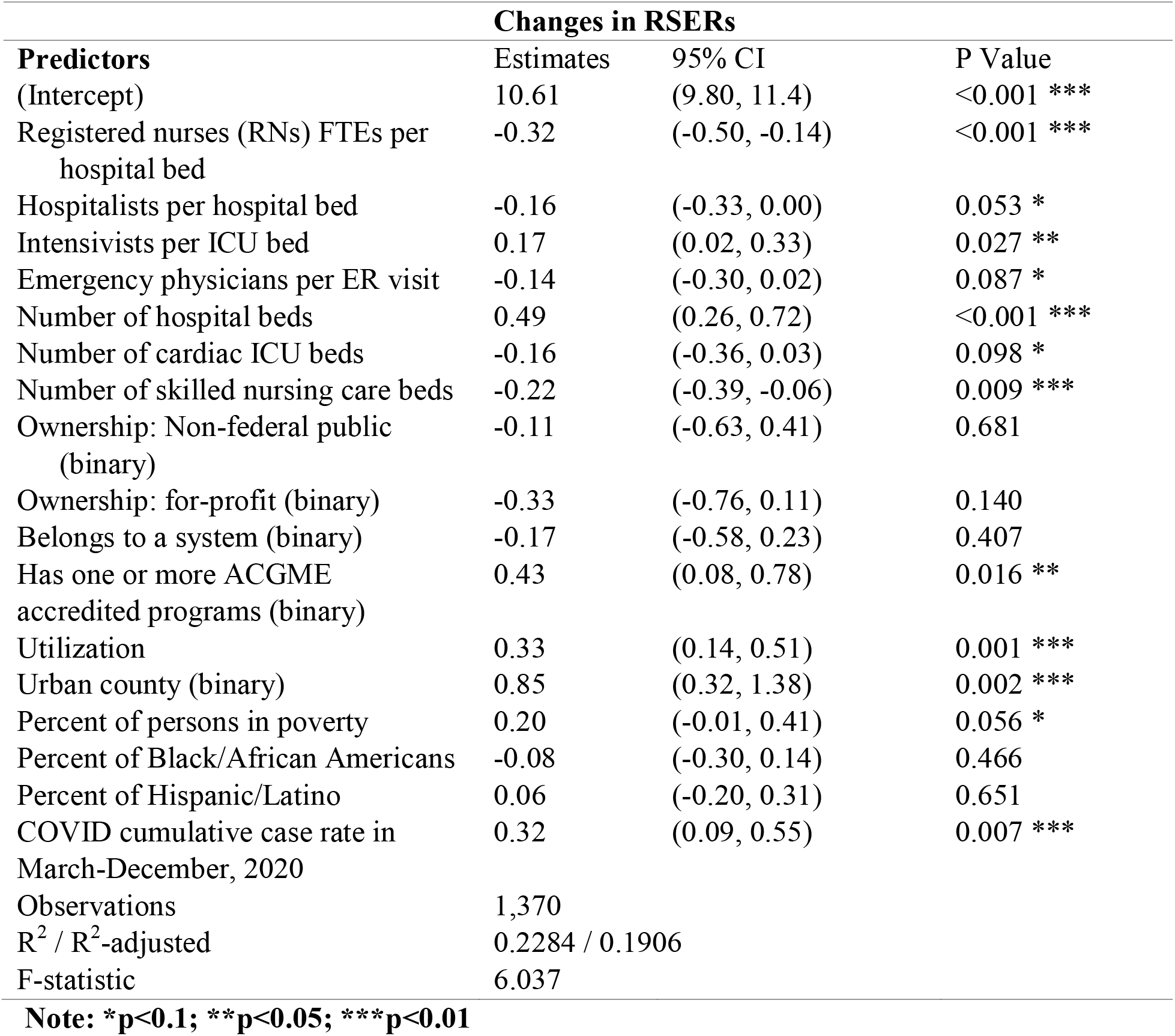
Parameter estimates quantifying the association between RSERs and hospital-and-county level attributes. Reported are the point estimates, 95% CIs, and p-values with superscripts highlighting statistical significance at 1%, 5%, and 10%. All continuous independent variables are centered and scaled.

Hospital baseline utilization is positively correlated with COVID-19 mortality rate, which indicates that hospitals with higher occupancy rates (estimate, 0.33; 95% CI, 0.14–0.51) tend to have worse patient outcome in terms of survival. Our regression results do not suggest any difference in mortality rate among not-for-profit, for-profit, and non-federal public hospitals. Whether or not a hospital belongs to a system is also not a significant predictor of mortality rate. Hospitals having one or more ACGME accredited program (estimate, 0.43; 95% CI, 0.08–0.78) tend to have higher mortality rates compared to hospitals that do not have such programs. At the county level, urban counties have higher COVID-19 mortality rates than rural counties (estimate, 0.85; 95% CI, 0.32–1.38). Percent of people in poverty (estimate, 0.20; 95% CI, -0.01–0.41) and cumulative COVID case rate (estimate, 0.32; 95% CI, 0.09–0.55) are positively associated with mortality rate. Conditionally on considering the other county level variables, percent of Black/African American residents and percent of Hispanic/Latino residents are not found to be statistically significant predictors in our data.

## Discussion

Traditional disaster planning in the United States focuses on the surge experienced by emergency departments during disasters.^22^ However, during pandemics, such as the current COVID-19 pandemic, general/surgical units, and ICUs are also overwhelmed. Therefore, it is imperative to examine how staffing levels across the hospital impact patient outcomes. This study relied on staffing levels from 2019, knowing that hospitals during this pandemic utilized their existing workforce to care for COVID-19 patients. While increased demand can overwhelm hospitals, bringing in additional providers from outside the hospital poses risks, including spreading the virus to hospital staff, and therefore was limited.^23^ Based on the regression results, COVID-19 patients’ receiving treatment in larger hospitals with lower RN, ER, and hospitalist staffing levels had the worst outcomes. Hospitals with more ICU beds and skilled nursing care beds had lower mortality rates. Moreover, hospitals with one or more ACGME accredited programs and hospitals with higher occupancy had higher COVID-19 mortality rates. In terms of county characteristics, hospitals located in urban areas and counties with higher COVID-19 infection rates had a lower likelihood of surviving.

Staffing levels matter! The positive relationship between staffing levels and patient outcomes has been documented for both RNs and hospitalists.^15,24,25^ Previous research examined the relationship between nurse staffing levels and COVID-19 infection and mortality rates in nursing home settings but not in hospital settings. Harrington et al. found a negative association between nurse staffing levels and infection rates in nursing homes.^26^ In another study, total nursing hours were associated with fewer deaths and a lower likelihood that the nursing home would experience an outbreak.^27^ Our study extends the literature to include hospital settings. Based on our statistical analysis, higher RN staffing levels are associated with lower inpatient COVID-19 mortality rates. Hospital administrators need to evaluate the heavy workload RNs experience during pandemics. Staffing levels and consequently workload not only predict patient outcomes during pandemics but has a direct impact on healthcare workers well-being.^28^ Ensuring adequate RN staffing levels protects not only the patients but also the well-being of our nursing workforce. It also eases the psychological stress RNs experience when simultaneously dealing with a surge in demand and the fear of carrying an infectious disease back home to their families and friends.^29^

Patients with severe COVID-19 infections are admitted to hospitals through the ER, where they are first treated and triaged by ER physicians. Afterward, if the patient is admitted, they are taken care by hospitalists and, in the case of ICU patients, by intensivists. ER physicians, particularly, are critical in the fight against morbidity and mortality from the novel coronavirus since they are the first point of contact. As Gaeta and Brennessel argue “physicians serve as a unique resource connecting a diverse patient population with emergent management of conditions.”^30^ Emergency department staff are responsible for triaging patients and for managing those patients with and without confirmed infection, all while being at the risk of contracting and spreading the virus and feeling burned out because of the long work shifts.^31^ This study found a (marginal) statistically significant negative relationship between emergency medicine staffing levels and COVID-19 mortality rates. This finding is important since there is limited research on the association between emergency medicine physicians and patient outcomes under normal circumstances and during pandemics.

Based on our study, hospitalists are another group of physicians whose staffing levels are negatively associated with COVID-19 mortality rates. As Yetmar et al. argue, hospitalists have the challenging task of “adapting to the many logistical and social elements of a pandemic.”^32^ Hospitalists’ role is made even more difficult since they are in direct contact with hospitalized COVID-19 patients and are at a higher risk of getting infected.^32^ Therefore, hospitals with higher hospitalist staffing levels are more prepared for the periods when a portion of the medical team gets sick and are more capable of meeting patient needs during surges in demand.

An interesting finding in this study is that higher intensivist staffing levels are associated with higher COVID-19 mortality rates. The number of ICU beds in the U.S. has been increasing since the inception of ICUs in the 1950s.^33^ Research on the relationship between intensivists staffing levels and patient outcomes is inconclusive. Dara and Afessa, for instance, compared mortality rates of patients admitted to the ICU based on intensivists-to-ICU beds ratio but found no significant difference in mortality rates.^34^ While Gershengorn et al. found that the relationship between intensivists staffing levels and mortality rates followed a U-shaped relationship.^35^ Lower levels and as well as high staffing levels were associated with higher mortality rates. It is important to note that there are different models of intensivist staffing. Some hospitals require the complete transfer of ICU patients to the care of a specific intensivists team, i.e. closed ICU model.^36^ In other hospitals, the responsibility of care for a patient is not transferred, but rather a consultation with an intensivist is required.^36^ Our study does not differentiate between these two models.

In terms of hospital beds, hospitals with a higher number of ICU and skilled nursing care beds had better outcomes, while hospitals with more surgical/medical beds had higher COVID-19 mortality rates. This finding is not surprising given that an adequate supply of ICU beds is needed to ensure that patients receive critical care in a timely manner. This is especially true during this pandemic whereby it is estimated that 16% of patients hospitalized for COVID-19 were admitted to the ICU.^37^ As De Lange et al. explain “lack of intensive care capacity has undoubtedly cost lives during the pandemic and will do again without greater baseline ICU capacity.”^38^ Skilled nursing beds might have helped hospitals during this pandemic by providing additional capacity to handle the surge. Alternatively, hospitals with skilled nursing beds might have been able to transfer COVID-19 patients who required rehabilitation more quickly out of the ICU. This would help make beds available in the ICU for newly admitted COVID-19 patients.

County-level variables that were associated with higher mortality rates were COVID-19 infection rates and urban setting. Previous research supports our finding that urban counties which tend to have higher population density and larger household size have higher mortality rates. Chandra et al., for instance, found a positive relationship between population density and mortality from the influenza pandemic of 1918-1919. ^39^ Tamblyn argue that communities with high population densities are more likely to experience outbreaks.^40^ Hospitals in counties with higher COVID-19 infections experience more severe demand surges which strains the local healthcare system and thus experience higher mortality rates.

## Conclusion

This study is not without limitations. Our sample is limited to patients whose outcomes are reported in the UnitedHealth Group Clinical Discovery database which, for example, specifically does not include those patients who are insured via Medicaid. Moreover, since we do not have reliable race data for the commercially insured patient population, we used residential level information to determine the distribution of patients by race at the hospital level. In addition, we rely on baseline data from 2019 to analyze hospital-level variables. Nevertheless, our study is the first to examine the association between RN and physician staffing levels with hospital-level COVID-19 mortality and includes a large geographically diverse cohort over a significant time span. Based on our findings, staffing levels are key predictors of patient outcomes. While policymakers have focused on increasing bed capacity, the supply of personal protective equipment, and ventilators, emergency preparedness plans and future policies should also take into account RN and hospital-based physician staffing levels. A future assessment is needed to establish benchmarks on the minimum staffing levels during pandemics. Based on these benchmarks, policies and plans should be developed to handle any shortage of hospital-based physicians during pandemics, given the impact staffing levels have on patient outcomes.

## Data Availability

All data produced in the present study are to remain private due to patient privacy concerns.

