## Supplemental Materials for "Inpatient COVID-19 Mortality Rates: What are the predictors?"

This supplemental material provides additional information about the methodologies, technical details, results, analytical dataset, and data definitions.

### A. Technical details

Let $i$ denote hospital and $j$ hospitalized patient associated with COVID diagnosis such that $i = 1, \ldots, I$ and $j=1,\ldots,n_{i}$, where $I$ is the total number of hospitals and $n_{i}$ is the total number of patients with COVID-related hospitalization. Let N = $\sum_{i=1}^{I} n_{i} be the total number of patients.$Let$Y_{ij}$ refer to the status of the $j$-th patient taking a value either 1 if a patient is deceased or transferred to hospice within 30-day of hospital admission date, or 0 otherwise. Denote by $\mathcal{X}_{ijk}$ the $k$-th covariate (e.g., gender, age, status of transfer from a nursing facility and Elixhauser comorbidities, discretized bins of time-differences); here $k = 1, \ldots, K$ with$K$ denoting the total number covariates.

We used a univariate association filter approach to select influential features. We fitted a series of univariate generalized linear models (GLM) with a logit link function corresponding to each variable of interest. We performed hypothesis tests for null effects via Wald test statistics, compared p-values, and selected the ones that are less than a prespecified threshold of 0.10. Elixhauser based comorbidities such as liver disease, peptic ulcer, and rheumatoid arthritis are excluded as the corresponding p-values were greater than 0.10. Alternatively, a more formal procedure like purposeful variable selection or stepwise variable selection can also be adopted. Numerical variables such as age and time differences between hospital admissions and March 1, 2020, are categorized into bins for ease of interpretability throughout the study. With an abuse of notation, denote by $X_{ijk}$ the $k$-th candidate covariate for the multivariate analysis that is to be discussed next; i.e$. X\subseteq\mathcal{X.}$

Hospital specific risk standardized event rates are estimated using a hierarchical model (which is also known as generalized linear mixed model (GLMM)) using the procedures described in Asch, et al.9 A fixed effect for logarithm of hospitals’ volume (i.e., average number of patients admitted in 2018-19) is added to account for potential variability in patient case-mix distribution among different sized hospitals. Alternatively, the volume of hospitals can represent the total number of COVID admitted patients in the sample; we observe moderate correlation (Pearson correlation coefficient 0.45 with 95% confidence interval (0.41, 0.49), p-value < 0.001) between these two measures. A multivariable GLMM framework, as below, is fitted

$${logit Pr(Y}_{ij}=1)=\beta_{0}+\sum_{k=1}^{K} X_{ijk}\beta_{k} + \gamma log({vol}_{i})+ b_{i0}\text{.}$$

Here fixed effect parameters, denoted by $\boldsymbol{\beta}= {(\beta}_{0}, \beta_{1},\ldots,\beta_{K},\gamma)$, quantify the effects of covariates on likelihood of experiencing events and characterize the deviation from the overall mean effect. Random effects $b_{i0}$ are assumed to follow Gaussian distribution with mean $0$and unknown variance $\sigma_{0b}^{2}$. From a hierarchical point-of-view, this model can be expressed as below

$${logit Pr(Y}_{ij}=1)= \mu_{i}+\sum_{k=1}^{K} X_{ijk}\beta_{k}\text{,}$$

$$\mu_{i}= \beta_{0}+\gamma log\left( {vol}_{i} \right)+ b_{i0}.$$

where $\mu_{i}$ represents hospital-specific mean.

Using the similar intuition to recycled predictions, RSERs are computed by averaging the event rates for all $N$ = $\sum_{i=1}^{I} n_{i}$ had each one of them been treated hypothetically in every hospital. We stack predicted probabilities in an $N\times I$ dimensional matrix and take column-wise average to obtain $I$ RSERs. Denote by $\tau$ a different hospital from $i$. Define the probability of experiencing the event if the $j$-th patient is coming from the $\tau$-th hospital

${p_{\tau j}=E(Y}_{\tau j}=1\left| X_{ij},{{vol}_{\tau}, b}_{\tau0} \right) = 1/\{1 + exp$(-$\beta_{0}-\sum_{k=1}^{K} X_{ijk}\beta_{k} - {\gamma log\left( {vol}_{\tau} \right)-b}_{\tau0})\}$.

Next, RSER for the $i$-th hospital is computed as

$$s_{i,DS}\text{ = }\sum_{\tau=1}^{I} \sum_{j=1}^{n_{\tau}} p_{\tau j} / N.$$

Such recycled event rates are also known as standardized event rates and entail probabilistic attribute.

Fixed and random effects are estimated using restricted maximum likelihood (REML) approach where the likelihood function is evaluated via adaptive Gauss-Hermite quadrature with 11 quadrature points and implemented by $GLMMadaptive$ R-package with default control parameters. The random effects are obtained as best linear unbiased predictors (BLUP) using a Bayesian formulation. We assessed the collinearity between variables via generalized variance inflation factor (GVIF) where the estimated GVIF are less than 2.00. We also fitted a multivariable GLM without random effects to check the appropriateness of using such random terms; the BIC are 61605.24 and 60810.17, and AIC are 61188.51 and 60574.27, respectively. REML estimate for the variance parameter associated with $b_{i0}$ is 0.22 with 95% CI (0.19, 0.26). Pearson’s correlation coefficient between the fixed effect estimated by GLM and GLMM is 1.00 (p-value < 0.001). The corresponding measures for the goodness-of-fit are conditional $R$-squared value (0.25), marginal R-squared (0.20), $C$-statistic (0.72), and Somer’s $D_{xy}$ (0.44).

### B. Results

Figure SM1 illustrates the adjusted odds ratios (aORs) with 95% confidence intervals (CIs) of prognostic risk factors that were treated as fixed effects in the hierarchical model. Figure SM2 exhibits the risk standardized event rates (RSERs) in ascending order for 1,397 hospitals estimated via recycled predictions – the higher the value, the worse the hospital performance is. Figure SM3 compares prevalence of different risk factors across two dominant pandemic waves (i.e., March 1 – May 31, 2020 vs October 1 – December 31, 2020); here the analytical dataset is a subset of the original dataset including sites having at least five patients in each wave resulting in a cohort of 43,858 patients distributed across 858 hospitals. Figure SM4 depicts the association between the ranks of same hospital (among 1,397 hospitals) computed from the main analytical dataset and sensitivity dataset. The latter relaxes the assumption of multiple hospital transfers; see Figure SM5.

Table SM1 details the data definitions and provides the corresponding ICD-10 codes. Table SM2 provides summary statistics of patient-level data for the main analytical dataset.

**Figure SM1.** Hierarchical model based adjusted odds ratios with 95% confidence intervals for risk factors; results are based on 95,919 patients admitted across 1,397 hospitals having at-least ten patients per site.

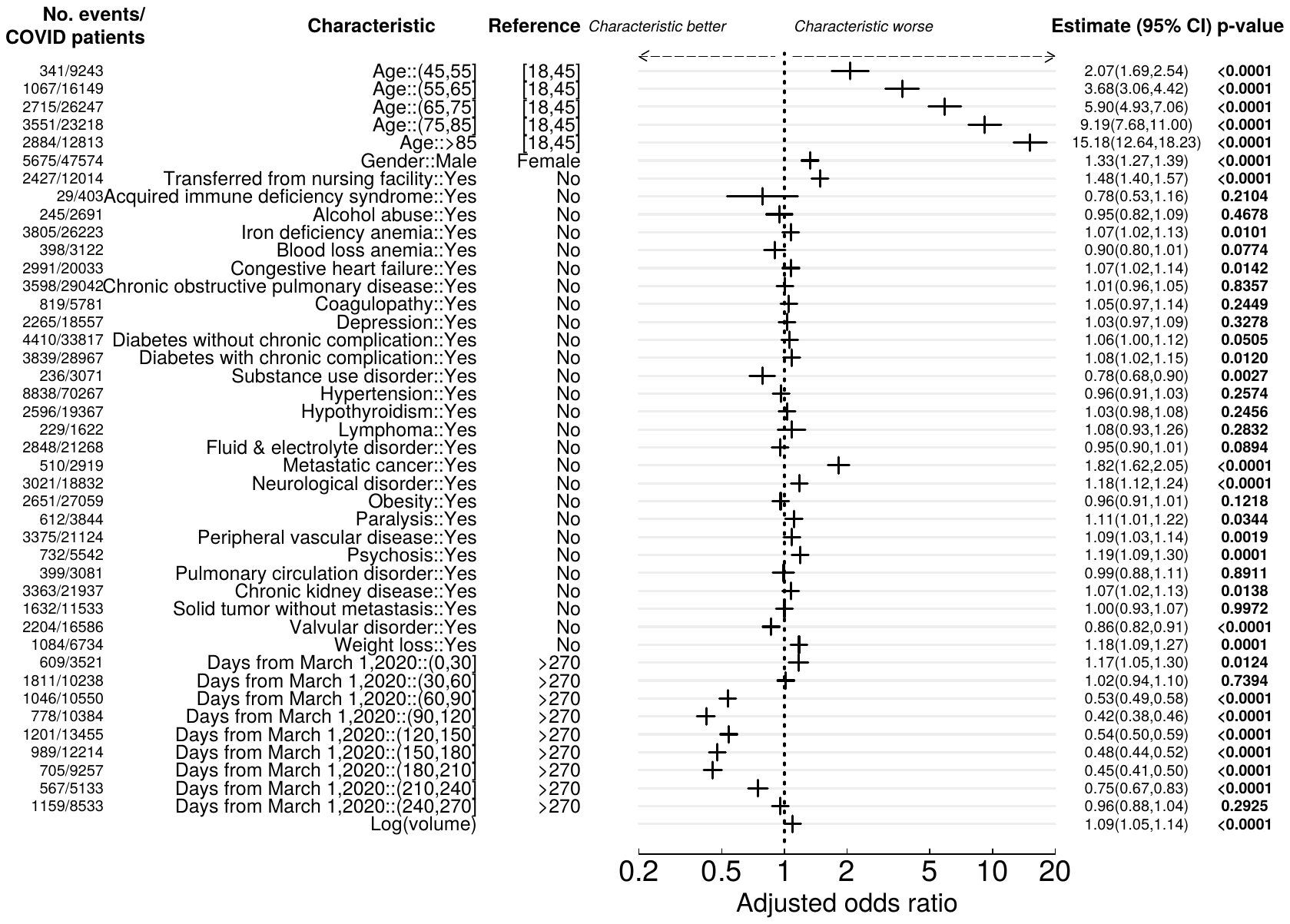

**Figure SM2**. Estimated RSERs for 1,397 hospitals; shaded area corresponds to inter quartile range (IQR) and dashed horizontal line refers to the observed event rate. Blue-colored “+” symbols represent hospitals with RSERs less than or equal to the observed event rate; and golden-colored “+” symbols highlight sites with RSERs greater than that.

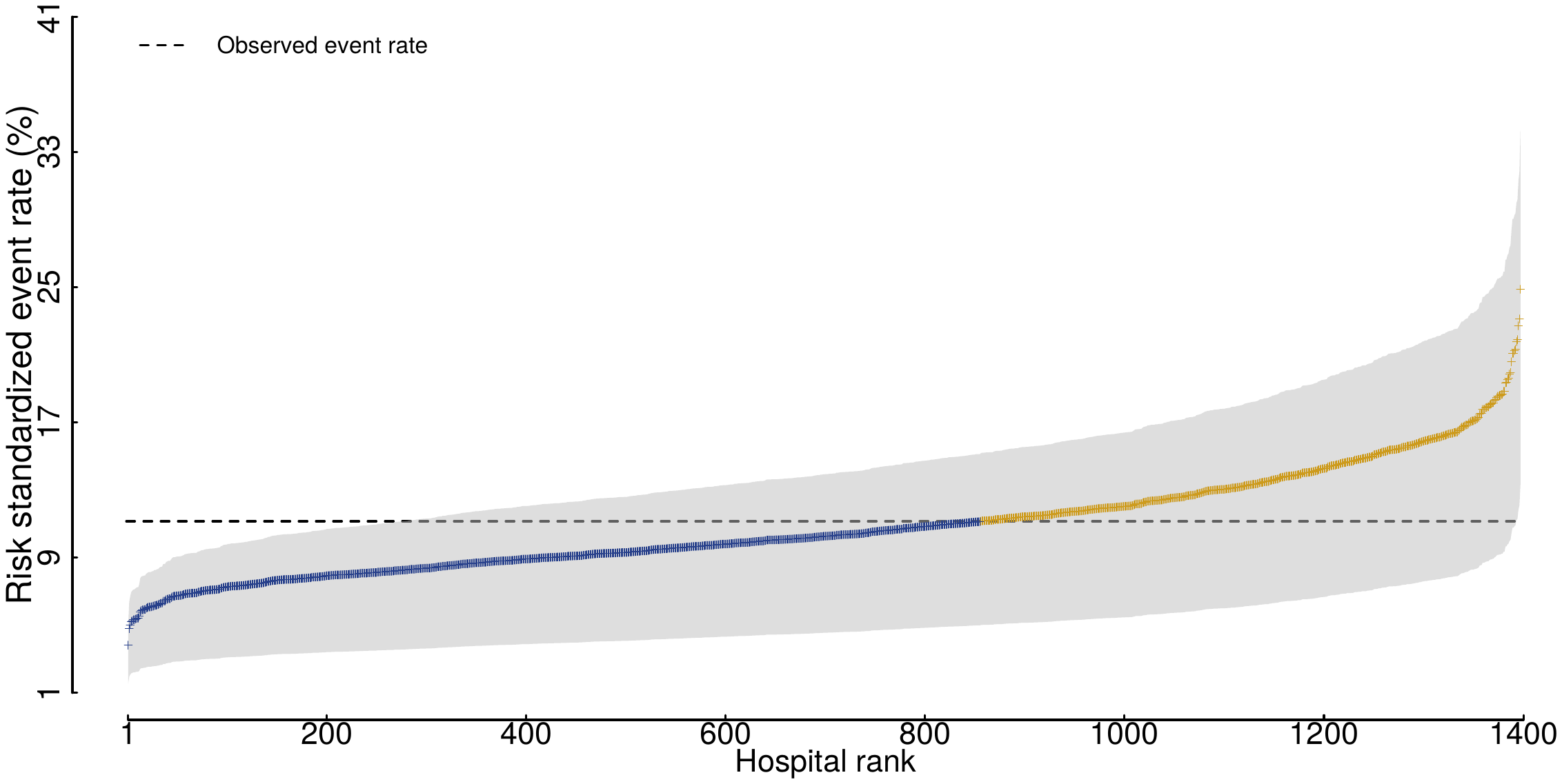

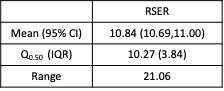

**Figure SM3**. Left panel shows the prevalence of risk factors between early (March 1-May 31, 2020) and late (October 1-Dcemeber 31) surges in Y-axis and X-axis respectively. Right panel exhibits the counts. Patients with more comorbidity were seen in the early surge comparing to the late surge.

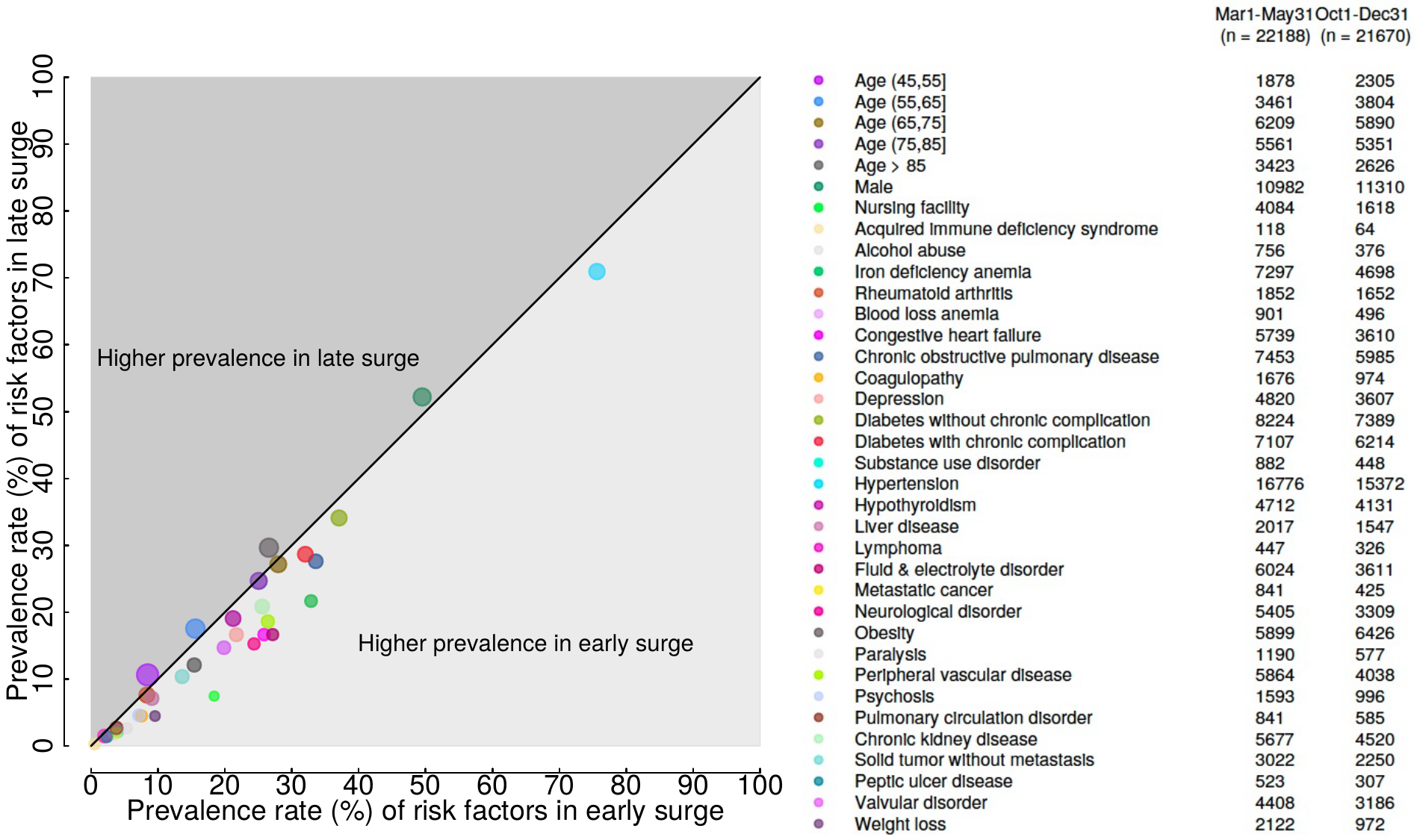

**Figure SM4.** Association between RSERs computed from the models based on original and sensitivity dataset. Rank correlation coefficient is reported for 1,397 sites’ RSERs based on two dataset.

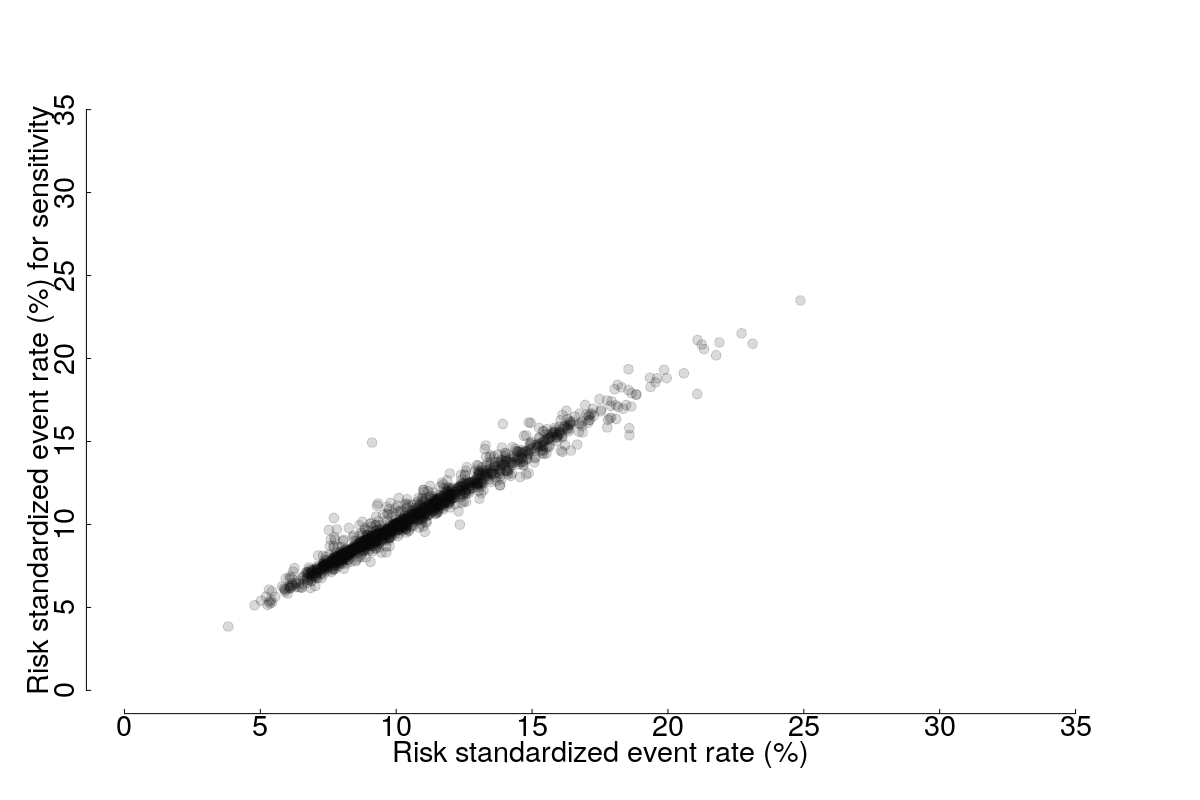

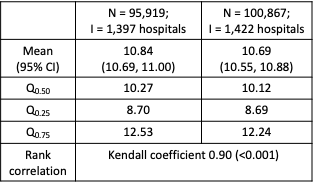

**Figure SM5.** Waterfall counts for the study datasets used for phase-1 and phase-2 analyses. Shaded boxes with blue color were used in phase-1 and phase-2 analyses; gray colored boxes were used for sensitivity analyses.

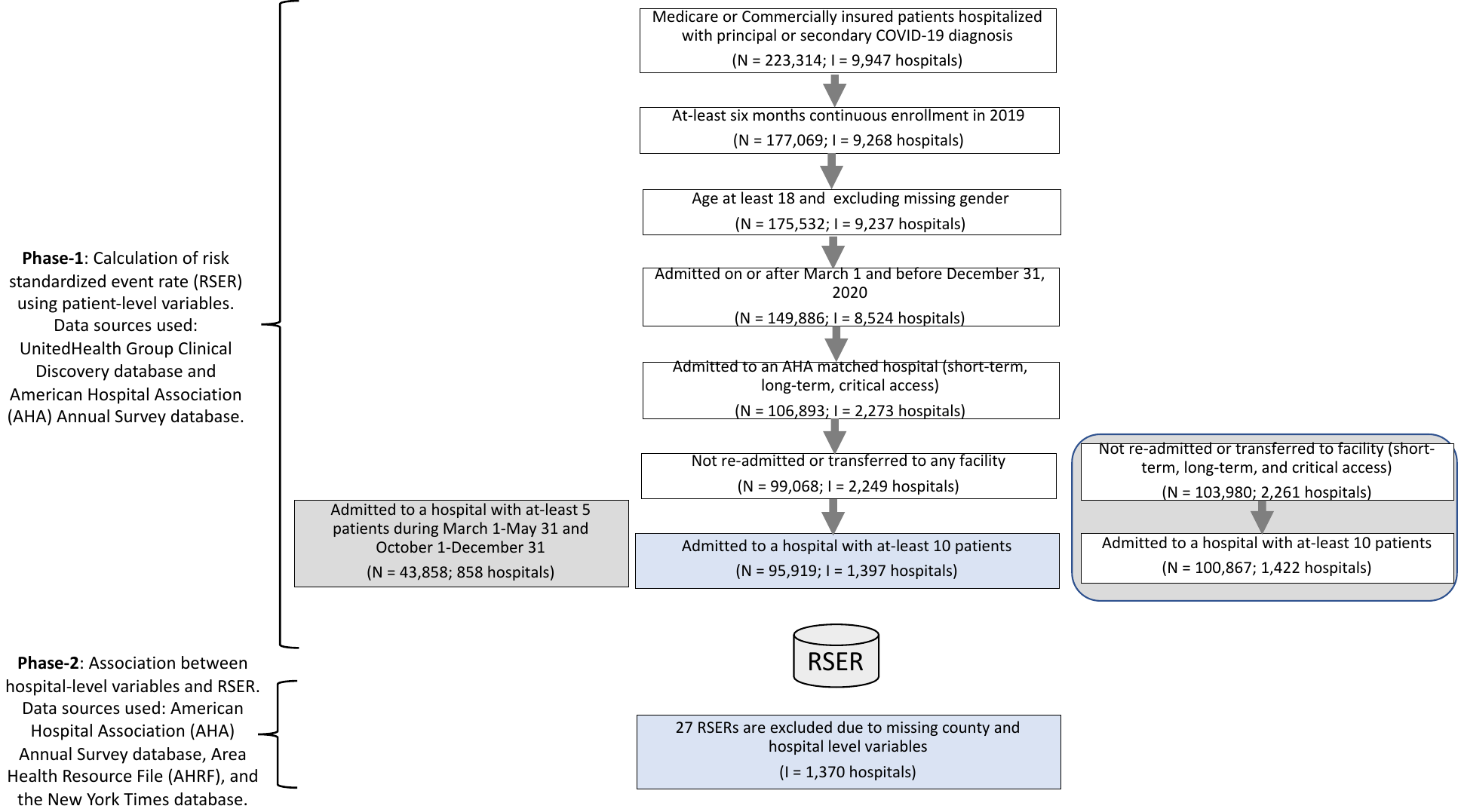

**Table SM1**: ICD-10 codes used to define variables.

**Inclusion Criteria for COVID diagnosis**

| Inclusion Criteria | ICD-10-CM codes |
| --- | --- |
| COVID-19 | U071, U072, B9729 |

**Elixhauser Comorbidity Indices**

| Elixhauser Comorbidity Index | ICD-10-CM codes |
| --- | --- |
| Acquired immunodeficiency Syndrome | B20 |
| Alcohol Use Disorder | F1010, F1011, F10120, F10121, F10129, F1014, F10150, F10151, F10159, F10180, F10181, F10182, F10188, F1019, F1020, F1021, F10220, F10221, F10229, F10230, F10231, F10232, F10239, F1024, F10250, F10251, F10259, F1026, F1027, F10280, F10281, F10282, F10288, F1029, F10921, F1094, F10950, F10951, F10959, F1096, F1097, F10980, F10981, F10982, F10988, F1099 |
| Iron Deficiency Anemia | D501, D508, D509, D510, D511, D512, D513, D518, D519, D520, D521, D528, D529, D530, D531, D532, D538, D539, D630, D631, D638, D649 |
| Rheumatoid Arthritis | L900, L940, L941, L943, M0500, M05011, M05012, M05019, M05021, M05022, M05029, M05031, M05032, M05039, M05041, M05042, M05049, M05051, M05052, M05059, M05061, M05062, M05069, M05071, M05072, M05079, M0509, M0510, M05111, M05112, M05119, M05121, M05122, M05129, M05131, M05132, M05139, M05141, M05142, M05149, M05151, M05152, M05159, M05161, M05162, M05169, M05171, M05172, M05179, M0519, M0520, M05211, M05212, M05219, M05221, M05222, M05229, M05231, M05232, M05239, M05241, M05242, M05249, M05251, M05252, M05259, M05261, M05262, M05269, M05271, M05272, M05279, M0529, M0530, M05311, M05312, M05319, M05321, M05322, M05329, M05331, M05332, M05339, M05341, M05342, M05349, M05351, M05352, M05359, M05361, M05362, M05369, M05371, M05372, M05379, M0539, M0540, M05411, M05412, M05419, M05421, M05422, M05429, M05431, M05432, M05439, M05441, M05442, M05449, M05451, M05452, M05459, M05461, M05462, M05469, M05471, M05472, M05479, M0549, M0550, M05511, M05512, M05519, M05521, M05522, M05529, M05531, M05532, M05539, M05541, M05542, M05549, M05551, M05552, M05559, M05561, M05562, M05569, M05571, M05572, M05579, M0559, M0560, M05611, M05612, M05619, M05621, M05622, M05629, M05631, M05632, M05639, M05641, M05642, M05649, M05651, M05652, M05659, M05661, M05662, M05669, M05671, M05672, M05679, M0569, M0570, M05711, M05712, M05719, M05721, M05722, M05729, M05731, M05732, M05739, M05741, M05742, M05749, M05751, M05752, M05759, M05761, M05762, M05769, M05771, M05772, M05779, M0579, M0580, M05811, M05812, M05819, M05821, M05822, M05829, M05831, M05832, M05839, M05841, M05842, M05849, M05851, M05852, M05859, M05861, M05862, M05869, M05871, M05872, M05879, M0589, M059, M0600, M06011, M06012, M06019, M06021, M06022, M06029, M06031, M06032, M06039, M06041, M06042, M06049, M06051, M06052, M06059, M06061, M06062, M06069, M06071, M06072, M06079, M0608, M0609, M061, M0620, M06211, M06212, M06219, M06221, M06222, M06229, M06231, M06232, M06239, M06241, M06242, M06249, M06251, M06252, M06259, M06261, M06262, M06269, M06271, M06272, M06279, M0628, M0629, M0630, M06311, M06312, M06319, M06321, M06322, M06329, M06331, M06332, M06339, M06341, M06342, M06349, M06351, M06352, M06359, M06361, M06362, M06369, M06371, M06372, M06379, M0638, M0639, M064, M0680, M06811, M06812, M06819, M06821, M06822, M06829, M06831, M06832, M06839, M06841, M06842, M06849, M06851, M06852, M06859, M06861, M06862, M06869, M06871, M06872, M06879, M0688, M0689, M069, M0800, M08011, M08012, M08019, M08021, M08022, M08029, M08031, M08032, M08039, M08041, M08042, M08049, M08051, M08052, M08059, M08061, M08062, M08069, M08071, M08072, M08079, M0808, M0809, M081, M0820, M08211, M08212, M08219, M08221, M08222, M08229, M08231, M08232, M08239, M08241, M08242, M08249, M08251, M08252, M08259, M08261, M08262, M08269, M08271, M08272, M08279, M0828, M0829, M083, M0840, M08411, M08412, M08419, M08421, M08422, M08429, M08431, M08432, M08439, M08441, M08442, M08449, M08451, M08452, M08459, M08461, M08462, M08469, M08471, M08472, M08479, M0848, M0880, M08811, M08812, M08819, M08821, M08822, M08829, M08831, M08832, M08839, M08841, M08842, M08849, M08851, M08852, M08859, M08861, M08862, M08869, M08871, M08872, M08879, M0888, M0889, M0890, M08911, M08912, M08919, M08921, M08922, M08929, M08931, M08932, M08939, M08941, M08942, M08949, M08951, M08952, M08959, M08961, M08962, M08969, M08971, M08972, M08979, M0898, M0899, M1200, M12011, M12012, M12019, M12021, M12022, M12029, M12031, M12032, M12039, M12041, M12042, M12049, M12051, M12052, M12059, M12061, M12062, M12069, M12071, M12072, M12079, M1208, M1209, M320, M3210, M3211, M3212, M3213, M3214, M3215, M3219, M328, M329, M3300, M3301, M3302, M3303, M3309, M3310, M3311, M3312, M3313, M3319, M3320, M3321, M3322, M3329, M3390, M3391, M3392, M3393, M3399, M340, M341, M342, M3481, M3482, M3483, M3489, M349, M3500, M3501, M3502, M3503, M3504, M3509, M351, M353, M355, M358, M359, M360, M368, M450, M451, M452, M453, M454, M455, M456, M457, M458, M459, M4600, M4601, M4602, M4603, M4604, M4605, M4606, M4607, M4608, M4609, M461, M4650, M4651, M4652, M4653, M4654, M4655, M4656, M4657, M4658, M4659, M4680, M4681, M4682, M4683, M4684, M4685, M4686, M4687, M4688, M4689, M4690, M4691, M4692, M4693, M4694, M4695, M4696, M4697, M4698, M4699, M488X1, M488X2, M488X3, M488X4, M488X5, M488X6, M488X7, M488X8, M488X9, M4980, M4981, M4982, M4983, M4984, M4985, M4986, M4987, M4988, M4989 |
| Blood Loss Anemia | D500, O9081, O99011, O99012, O99013, O99019, O9902, O9903 |
| Congestive Heart Failure | I0981, I110, I130, I132, I501, I5020, I5021, I5022, I5023, I5030, I5031, I5032, I5033, I5040, I5041, I5042, I5043, I50810, I50811, I50812, I50813, I50814, I5082, I5083, I5084, I5089, I509 |
| Chronic Obstructive Pulmonary Disease | J40, J410, J411, J418, J42, J430, J431, J432, J438, J439, J440, J441, J449, J4520, J4521, J4522, J4530, J4531, J4532, J4540, J4541, J4542, J4550, J4551, J4552, J45901, J45902, J45909, J45990, J45991, J45998, J470, J471, J479, J60, J61, J620, J628, J630, J631, J632, J633, J634, J635, J636, J64, J660, J661, J662, J668, J670, J671, J672, J673, J674, J675, J676, J677, J678, J679, J684 |
| Coagulopathy | D65, D66, D67, D680, D681, D682, D68311, D68312, D68318, D6832, D684, D688, D689, D691, D693, D6941, D6942, D6949, D6951, D6959, D696, D7582, O99111, O99112, O99113, O99119, O9912, O9913 |
| Depression | F320, F321, F322, F323, F328, F3281, F3289, F329, F330, F331, F332, F333, F338, F339, F341, F4321 |
| Diabetes without Chronic Complications | E0800, E0801, E0810, E0811, E089, E0900, E0901, E0910, E0911, E099, E1010, E1011, E109, E1100, E1101, E1110, E1111, E119, E1300, E1301, E1310, E1311, E139, O24011, O24012, O24013, O24019, O2402, O2403, O24111, O24112, O24113, O24119, O2412, O2413, O24311, O24312, O24313, O24319, O2432, O2433, O24811, O24812, O24813, O24819, O2482, O2483, O24911, O24912, O24913, O24919, O2492, O2493 |
| Diabetes with Chronic Complications | E0821, E0822, E0829, E08311, E08319, E08321, E083211, E083212, E083213, E083219, E08329, E083291, E083292, E083293, E083299, E08331, E083311, E083312, E083313, E083319, E08339, E083391, E083392, E083393, E083399, E08341, E083411, E083412, E083413, E083419, E08349, E083491, E083492, E083493, E083499, E08351, E083511, E083512, E083513, E083519, E083521, E083522, E083523, E083529, E083531, E083532, E083533, E083539, E083541, E083542, E083543, E083549, E083551, E083552, E083553, E083559, E08359, E083591, E083592, E083593, E083599, E0836, E0837X1, E0837X2, E0837X3, E0837X9, E0839, E0840, E0841, E0842, E0843, E0844, E0849, E0851, E0852, E0859, E08610, E08618, E08620, E08621, E08622, E08628, E08630, E08638, E08641, E08649, E0865, E0869, E088, E0921, E0922, E0929, E09311, E09319, E09321, E093211, E093212, E093213, E093219, E09329, E093291, E093292, E093293, E093299, E09331, E093311, E093312, E093313, E093319, E09339, E093391, E093392, E093393, E093399, E09341, E093411, E093412, E093413, E093419, E09349, E093491, E093492, E093493, E093499, E09351, E093511, E093512, E093513, E093519, E093521, E093522, E093523, E093529, E093531, E093532, E093533, E093539, E093541, E093542, E093543, E093549, E093551, E093552, E093553, E093559, E09359, E093591, E093592, E093593, E093599, E0936, E0937X1, E0937X2, E0937X3, E0937X9, E0939, E0940, E0941, E0942, E0943, E0944, E0949, E0951, E0952, E0959, E09610, E09618, E09620, E09621, E09622, E09628, E09630, E09638, E09641, E09649, E0965, E0969, E098, E1021, E1022, E1029, E10311, E10319, E10321, E103211, E103212, E103213, E103219, E10329, E103291, E103292, E103293, E103299, E10331, E103311, E103312, E103313, E103319, E10339, E103391, E103392, E103393, E103399, E10341, E103411, E103412, E103413, E103419, E10349, E103491, E103492, E103493, E103499, E10351, E103511, E103512, E103513, E103519, E103521, E103522, E103523, E103529, E103531, E103532, E103533, E103539, E103541, E103542, E103543, E103549, E103551, E103552, E103553, E103559, E10359, E103591, E103592, E103593, E103599, E1036, E1037X1, E1037X2, E1037X3, E1037X9, E1039, E1040, E1041, E1042, E1043, E1044, E1049, E1051, E1052, E1059, E10610, E10618, E10620, E10621, E10622, E10628, E10630, E10638, E10641, E10649, E1065, E1069, E108, E1121, E1122, E1129, E11311, E11319, E11321, E113211, E113212, E113213, E113219, E11329, E113291, E113292, E113293, E113299, E11331, E113311, E113312, E113313, E113319, E11339, E113391, E113392, E113393, E113399, E11341, E113411, E113412, E113413, E113419, E11349, E113491, E113492, E113493, E113499, E11351, E113511, E113512, E113513, E113519, E113521, E113522, E113523, E113529, E113531, E113532, E113533, E113539, E113541, E113542, E113543, E113549, E113551, E113552, E113553, E113559, E11359, E113591, E113592, E113593, E113599, E1136, E1137X1, E1137X2, E1137X3, E1137X9, E1139, E1140, E1141, E1142, E1143, E1144, E1149, E1151, E1152, E1159, E11610, E11618, E11620, E11621, E11622, E11628, E11630, E11638, E11641, E11649, E1165, E1169, E118, E1321, E1322, E1329, E13311, E13319, E13321, E133211, E133212, E133213, E133219, E13329, E133291, E133292, E133293, E133299, E13331, E133311, E133312, E133313, E133319, E13339, E133391, E133392, E133393, E133399, E13341, E133411, E133412, E133413, E133419, E13349, E133491, E133492, E133493, E133499, E13351, E133511, E133512, E133513, E133519, E133521, E133522, E133523, E133529, E133531, E133532, E133533, E133539, E133541, E133542, E133543, E133549, E133551, E133552, E133553, E133559, E13359, E133591, E133592, E133593, E133599, E1336, E1337X1, E1337X2, E1337X3, E1337X9, E1339, E1340, E1341, E1342, E1343, E1344, E1349, E1351, E1352, E1359, E13610, E13618, E13620, E13621, E13622, E13628, E13630, E13638, E13641, E13649, E1365, E1369, E138, P702 |
| Substance Use Disorder | F1110, F1111, F11120, F11121, F11122, F11129, F1114, F11150, F11151, F11159, F11181, F11182, F11188, F1119, F1120, F1121, F11220, F11221, F11222, F11229, F1123, F1124, F11250, F11251, F11259, F11281, F11282, F11288, F1129, F1210, F1211, F12120, F12121, F12122, F12129, F12150, F12151, F12159, F12180, F12188, F1219, F1220, F1221, F12220, F12221, F12222, F12229, F1223, F12250, F12251, F12259, F12280, F12288, F1229, F1310, F1311, F13120, F13121, F13129, F1314, F13150, F13151, F13159, F13180, F13181, F13182, F13188, F1319, F1320, F1321, F13220, F13221, F13229, F13230, F13231, F13232, F13239, F1324, F13250, F13251, F13259, F1326, F1327, F13280, F13281, F13282, F13288, F1329, F1410, F1411, F14120, F14121, F14122, F14129, F1414, F14150, F14151, F14159, F14180, F14181, F14182, F14188, F1419, F1420, F1421, F14220, F14221, F14222, F14229, F1423, F1424, F14250, F14251, F14259, F14280, F14281, F14282, F14288, F1429, F1510, F1511, F15120, F15121, F15122, F15129, F1514, F15150, F15151, F15159, F15180, F15181, F15182, F15188, F1519, F1520, F1521, F15220, F15221, F15222, F15229, F1523, F1524, F15250, F15251, F15259, F15280, F15281, F15282, F15288, F1529, F1610, F1611, F16120, F16121, F16122, F16129, F1614, F16150, F16151, F16159, F16180, F16183, F16188, F1619, F1620, F1621, F16220, F16221, F16229, F1624, F16250, F16251, F16259, F16280, F16283, F16288, F1629, F1810, F1811, F18120, F18121, F18129, F1814, F18150, F18151, F18159, F1817, F18180, F18188, F1819, F1820, F1821, F18220, F18221, F18229, F1824, F18250, F18251, F18259, F1827, F18280, F18288, F1829, F1910, F1911, F19120, F19121, F19122, F19129, F1914, F19150, F19151, F19159, F1916, F1917, F19180, F19181, F19182, F19188, F1919, F1920, F1921, F19220, F19221, F19222, F19229, F19230, F19231, F19232, F19239, F1924, F19250, F19251, F19259, F1926, F1927, F19280, F19281, F19282, F19288, F1929, F550, F551, F552, F553, F554, F558, O99320, O99321, O99322, O99323, O99324, O99325 |
| Hypertension | I10, I110, I119, I120, I129, I130, I1310, I1311, I132, I150, I151, I152, I158, I159, I160, I161, I169, I674, O10011, O10012, O10013, O10019, O1002, O1003, O10111, O10112, O10113, O10119, O1012, O1013, O10211, O10212, O10213, O10219, O1022, O1023, O10311, O10312, O10313, O10319, O1032, O1033, O10411, O10412, O10413, O10419, O1042, O1043, O10911, O10912, O10913, O10919, O1092, O1093, O111, O112, O113, O114, O115, O119, O161, O162, O163, O164, O165, O169 |
| Hypothyroidism | E000, E001, E002, E009, E018, E02, E030, E031, E032, E033, E038, E039, E890 |
| Liver Disease | B180, B181, B182, I8500, I8501, I8510, I8511, K700, K702, K7030, K7031, K7040, K7041, K709, K7210, K7211, K7290, K7291, K730, K731, K732, K738, K739, K740, K741, K742, K743, K744, K745, K7460, K7469, K754, K7581, K760, K766, K7689, K769, Z944 |
| Lymphoma | C8100, C8101, C8102, C8103, C8104, C8105, C8106, C8107, C8108, C8109, C8110, C8111, C8112, C8113, C8114, C8115, C8116, C8117, C8118, C8119, C8120, C8121, C8122, C8123, C8124, C8125, C8126, C8127, C8128, C8129, C8130, C8131, C8132, C8133, C8134, C8135, C8136, C8137, C8138, C8139, C8140, C8141, C8142, C8143, C8144, C8145, C8146, C8147, C8148, C8149, C8170, C8171, C8172, C8173, C8174, C8175, C8176, C8177, C8178, C8179, C8190, C8191, C8192, C8193, C8194, C8195, C8196, C8197, C8198, C8199, C8200, C8201, C8202, C8203, C8204, C8205, C8206, C8207, C8208, C8209, C8210, C8211, C8212, C8213, C8214, C8215, C8216, C8217, C8218, C8219, C8220, C8221, C8222, C8223, C8224, C8225, C8226, C8227, C8228, C8229, C8230, C8231, C8232, C8233, C8234, C8235, C8236, C8237, C8238, C8239, C8240, C8241, C8242, C8243, C8244, C8245, C8246, C8247, C8248, C8249, C8250, C8251, C8252, C8253, C8254, C8255, C8256, C8257, C8258, C8259, C8260, C8261, C8262, C8263, C8264, C8265, C8266, C8267, C8268, C8269, C8280, C8281, C8282, C8283, C8284, C8285, C8286, C8287, C8288, C8289, C8290, C8291, C8292, C8293, C8294, C8295, C8296, C8297, C8298, C8299, C8300, C8301, C8302, C8303, C8304, C8305, C8306, C8307, C8308, C8309, C8310, C8311, C8312, C8313, C8314, C8315, C8316, C8317, C8318, C8319, C8330, C8331, C8332, C8333, C8334, C8335, C8336, C8337, C8338, C8339, C8350, C8351, C8352, C8353, C8354, C8355, C8356, C8357, C8358, C8359, C8370, C8371, C8372, C8373, C8374, C8375, C8376, C8377, C8378, C8379, C8380, C8381, C8382, C8383, C8384, C8385, C8386, C8387, C8388, C8389, C8390, C8391, C8392, C8393, C8394, C8395, C8396, C8397, C8398, C8399, C8400, C8401, C8402, C8403, C8404, C8405, C8406, C8407, C8408, C8409, C8410, C8411, C8412, C8413, C8414, C8415, C8416, C8417, C8418, C8419, C8440, C8441, C8442, C8443, C8444, C8445, C8446, C8447, C8448, C8449, C8460, C8461, C8462, C8463, C8464, C8465, C8466, C8467, C8468, C8469, C8470, C8471, C8472, C8473, C8474, C8475, C8476, C8477, C8478, C8479, C8490, C8491, C8492, C8493, C8494, C8495, C8496, C8497, C8498, C8499, C84A0, C84A1, C84A2, C84A3, C84A4, C84A5, C84A6, C84A7, C84A8, C84A9, C84Z0, C84Z1, C84Z2, C84Z3, C84Z4, C84Z5, C84Z6, C84Z7, C84Z8, C84Z9, C8510, C8511, C8512, C8513, C8514, C8515, C8516, C8517, C8518, C8519, C8520, C8521, C8522, C8523, C8524, C8525, C8526, C8527, C8528, C8529, C8580, C8581, C8582, C8583, C8584, C8585, C8586, C8587, C8588, C8589, C8590, C8591, C8592, C8593, C8594, C8595, C8596, C8597, C8598, C8599, C860, C861, C862, C863, C864, C865, C866, C880, C882, C883, C884, C888, C889, C9000, C9001, C9002, C9010, C9011, C9012, C9020, C9021, C9022, C9030, C9031, C9032, C960, C962, C9620, C9621, C9622, C9629, C964, C969, C96A, C96Z, D47Z9 |
| Fluid and Electrolyte Disorder | E860, E861, E869, E870, E871, E872, E873, E874, E875, E876, E8770, E8771, E8779, E878 |
| Metastatic Cancer | C770, C771, C772, C773, C774, C775, C778, C779, C7800, C7801, C7802, C781, C782, C7830, C7839, C784, C785, C786, C787, C7880, C7889, C7900, C7901, C7902, C7910, C7911, C7919, C792, C7931, C7932, C7940, C7949, C7951, C7952, C7960, C7961, C7962, C7970, C7971, C7972, C7981, C7982, C7989, C799, C7B00, C7B01, C7B02, C7B03, C7B04, C7B09, C7B1, C7B8, C800, C801, R180 |
| Neurological Disorder | E7500, E7501, E7502, E7509, E7510, E7511, E7519, E7523, E7525, E7526, E7529, E754, F842, G10, G110, G111, G112, G113, G114, G118, G119, G120, G121, G1220, G1221, G1222, G1223, G1224, G1225, G1229, G128, G129, G132, G138, G20, G214, G2401, G2402, G2409, G242, G248, G254, G255, G2581, G300, G301, G308, G309, G3101, G3109, G311, G312, G3181, G3182, G3183, G3184, G3185, G3189, G319, G3281, G35, G361, G368, G369, G370, G371, G372, G373, G374, G375, G378, G379, G40001, G40009, G40011, G40019, G40101, G40109, G40111, G40119, G40201, G40209, G40211, G40219, G40301, G40309, G40311, G40319, G40401, G40409, G40411, G40419, G40501, G40509, G40801, G40802, G40803, G40804, G40811, G40812, G40813, G40814, G40821, G40822, G40823, G40824, G4089, G40901, G40909, G40911, G40919, G40A01, G40A09, G40A11, G40A19, G40B01, G40B09, G40B11, G40B19, G47411, G47419, G47421, G47429, G803, G890, G910, G911, G912, G913, G914, G918, G919, G937, G9389, G939, G94, O99350, O99351, O99352, O99353, O99354, O99355, P9160, P9161, P9162, P9163, R410, R4182, R4701, R5600, R5601, R561, R569 |
| Obesity | E6601, E6609, E661, E662, E668, E669, O99210, O99211, O99212, O99213, O99214, O99215, R939, Z6830, Z6831, Z6832, Z6833, Z6834, Z6835, Z6836, Z6837, Z6838, Z6839, Z6841, Z6842, Z6843, Z6844, Z6845, Z6854 |
| Paralysis | G041, G800, G801, G802, G804, G808, G809, G8100, G8101, G8102, G8103, G8104, G8110, G8111, G8112, G8113, G8114, G8190, G8191, G8192, G8193, G8194, G8220, G8221, G8222, G8250, G8251, G8252, G8253, G8254, G830, G8310, G8311, G8312, G8313, G8314, G8320, G8321, G8322, G8323, G8324, G8330, G8331, G8332, G8333, G8334, G834, G835, G8381, G8382, G8383, G8384, G8389, G839, I69031, I69032, I69033, I69034, I69039, I69041, I69042, I69043, I69044, I69049, I69051, I69052, I69053, I69054, I69059, I69061, I69062, I69063, I69064, I69065, I69069, I69131, I69132, I69133, I69134, I69139, I69141, I69142, I69143, I69144, I69149, I69151, I69152, I69153, I69154, I69159, I69161, I69162, I69163, I69164, I69165, I69169, I69231, I69232, I69233, I69234, I69239, I69241, I69242, I69243, I69244, I69249, I69251, I69252, I69253, I69254, I69259, I69261, I69262, I69263, I69264, I69265, I69269, I69331, I69332, I69333, I69334, I69339, I69341, I69342, I69343, I69344, I69349, I69351, I69352, I69353, I69354, I69359, I69361, I69362, I69363, I69364, I69365, I69369, I69831, I69832, I69833, I69834, I69839, I69841, I69842, I69843, I69844, I69849, I69851, I69852, I69853, I69854, I69859, I69861, I69862, I69863, I69864, I69865, I69869, I69931, I69932, I69933, I69934, I69939, I69941, I69942, I69943, I69944, I69949, I69951, I69952, I69953, I69954, I69959, I69961, I69962, I69963, I69964, I69965, I69969, R532 |
| Peripheral Vascular Disease | I700, I701, I70201, I70202, I70203, I70208, I70209, I70211, I70212, I70213, I70218, I70219, I70221, I70222, I70223, I70228, I70229, I70231, I70232, I70233, I70234, I70235, I70238, I70239, I70241, I70242, I70243, I70244, I70245, I70248, I70249, I7025, I70261, I70262, I70263, I70268, I70269, I70291, I70292, I70293, I70298, I70299, I70301, I70302, I70303, I70308, I70309, I70311, I70312, I70313, I70318, I70319, I70321, I70322, I70323, I70328, I70329, I70331, I70332, I70333, I70334, I70335, I70338, I70339, I70341, I70342, I70343, I70344, I70345, I70348, I70349, I7035, I70361, I70362, I70363, I70368, I70369, I70391, I70392, I70393, I70398, I70399, I70401, I70402, I70403, I70408, I70409, I70411, I70412, I70413, I70418, I70419, I70421, I70422, I70423, I70428, I70429, I70431, I70432, I70433, I70434, I70435, I70438, I70439, I70441, I70442, I70443, I70444, I70445, I70448, I70449, I7045, I70461, I70462, I70463, I70468, I70469, I70491, I70492, I70493, I70498, I70499, I70501, I70502, I70503, I70508, I70509, I70511, I70512, I70513, I70518, I70519, I70521, I70522, I70523, I70528, I70529, I70531, I70532, I70533, I70534, I70535, I70538, I70539, I70541, I70542, I70543, I70544, I70545, I70548, I70549, I7055, I70561, I70562, I70563, I70568, I70569, I70591, I70592, I70593, I70598, I70599, I70601, I70602, I70603, I70608, I70609, I70611, I70612, I70613, I70618, I70619, I70621, I70622, I70623, I70628, I70629, I70631, I70632, I70633, I70634, I70635, I70638, I70639, I70641, I70642, I70643, I70644, I70645, I70648, I70649, I7065, I70661, I70662, I70663, I70668, I70669, I70691, I70692, I70693, I70698, I70699, I70701, I70702, I70703, I70708, I70709, I70711, I70712, I70713, I70718, I70719, I70721, I70722, I70723, I70728, I70729, I70731, I70732, I70733, I70734, I70735, I70738, I70739, I70741, I70742, I70743, I70744, I70745, I70748, I70749, I7075, I70761, I70762, I70763, I70768, I70769, I70791, I70792, I70793, I70798, I70799, I708, I7090, I7091, I7092, I7100, I7101, I7102, I7103, I711, I712, I713, I714, I715, I716, I718, I719, I720, I721, I722, I723, I724, I725, I726, I728, I729, I731, I7381, I7389, I739, I742, I743, I744, I76, I771, I7770, I7771, I7772, I7773, I7774, I7775, I7776, I7777, I7779, I790, I791, I798, K551, K558, K559, Z95820, Z95828 |
| Psychosis | F200, F201, F202, F203, F205, F2081, F2089, F209, F22, F23, F24, F250, F251, F258, F259, F28, F29, F3010, F3011, F3012, F3013, F302, F303, F304, F308, F309, F310, F3110, F3111, F3112, F3113, F312, F3130, F3131, F3132, F314, F315, F3160, F3161, F3162, F3163, F3164, F3170, F3171, F3172, F3173, F3174, F3175, F3176, F3177, F3178, F3181, F3189, F319, F324, F325, F3340, F3341, F3342, F348, F3481, F3489, F349, F39, F4489, F843 |
| Pulmonary Circulation | I2601, I2602, I2609, I2690, I2692, I2699, I270, I271, I2781, I2782, I2783, I2789, I279, I289, T800XXA, T82817A, T82818A |
| Chronic Kidney Disease | I120, I1311, I132, N183, N184, N185, N186, N189, N19, Z4901, Z4902, Z4931, Z4932, Z9115, Z940, Z992 |
| Solid Tumor without Metastasis | C000, C001, C002, C003, C004, C005, C006, C008, C009, C01, C020, C021, C022, C023, C024, C028, C029, C030, C031, C039, C040, C041, C048, C049, C050, C051, C052, C058, C059, C060, C061, C062, C0680, C0689, C069, C07, C080, C081, C089, C090, C091, C098, C099, C100, C101, C102, C103, C104, C108, C109, C110, C111, C112, C113, C118, C119, C12, C130, C131, C132, C138, C139, C140, C142, C148, C153, C154, C155, C158, C159, C160, C161, C162, C163, C164, C165, C166, C168, C169, C170, C171, C172, C173, C178, C179, C180, C181, C182, C183, C184, C185, C186, C187, C188, C189, C19, C20, C210, C211, C212, C218, C220, C221, C222, C223, C224, C227, C228, C229, C23, C240, C241, C248, C249, C250, C251, C252, C253, C254, C257, C258, C259, C260, C261, C269, C300, C301, C310, C311, C312, C313, C318, C319, C320, C321, C322, C323, C328, C329, C33, C3400, C3401, C3402, C3410, C3411, C3412, C342, C3430, C3431, C3432, C3480, C3481, C3482, C3490, C3491, C3492, C37, C380, C381, C382, C383, C384, C388, C390, C399, C4000, C4001, C4002, C4010, C4011, C4012, C4020, C4021, C4022, C4030, C4031, C4032, C4080, C4081, C4082, C4090, C4091, C4092, C410, C411, C412, C413, C414, C419, C430, C4310, C4311, C43111, C43112, C4312, C43121, C43122, C4320, C4321, C4322, C4330, C4331, C4339, C434, C4351, C4352, C4359, C4360, C4361, C4362, C4370, C4371, C4372, C438, C439, C450, C451, C452, C457, C470, C4710, C4711, C4712, C4720, C4721, C4722, C473, C474, C475, C476, C478, C479, C480, C481, C482, C488, C490, C4910, C4911, C4912, C4920, C4921, C4922, C493, C494, C495, C496, C498, C499, C49A0, C49A1, C49A2, C49A3, C49A4, C49A5, C49A9, C4A0, C4A10, C4A11, C4A111, C4A112, C4A12, C4A121, C4A122, C4A20, C4A21, C4A22, C4A30, C4A31, C4A39, C4A4, C4A51, C4A52, C4A59, C4A60, C4A61, C4A62, C4A70, C4A71, C4A72, C4A8, C4A9, C50011, C50012, C50019, C50021, C50022, C50029, C50111, C50112, C50119, C50121, C50122, C50129, C50211, C50212, C50219, C50221, C50222, C50229, C50311, C50312, C50319, C50321, C50322, C50329, C50411, C50412, C50419, C50421, C50422, C50429, C50511, C50512, C50519, C50521, C50522, C50529, C50611, C50612, C50619, C50621, C50622, C50629, C50811, C50812, C50819, C50821, C50822, C50829, C50911, C50912, C50919, C50921, C50922, C50929, C510, C511, C512, C518, C519, C52, C530, C531, C538, C539, C540, C541, C542, C543, C548, C549, C55, C561, C562, C569, C5700, C5701, C5702, C5710, C5711, C5712, C5720, C5721, C5722, C573, C574, C577, C578, C579, C58, C600, C601, C602, C608, C609, C61, C6200, C6201, C6202, C6210, C6211, C6212, C6290, C6291, C6292, C6300, C6301, C6302, C6310, C6311, C6312, C632, C637, C638, C639, C641, C642, C649, C651, C652, C659, C661, C662, C669, C670, C671, C672, C673, C674, C675, C676, C677, C678, C679, C680, C681, C688, C689, C6900, C6901, C6902, C6910, C6911, C6912, C6920, C6921, C6922, C6930, C6931, C6932, C6940, C6941, C6942, C6950, C6951, C6952, C6960, C6961, C6962, C6980, C6981, C6982, C6990, C6991, C6992, C700, C701, C709, C710, C711, C712, C713, C714, C715, C716, C717, C718, C719, C720, C721, C7220, C7221, C7222, C7230, C7231, C7232, C7240, C7241, C7242, C7250, C7259, C729, C73, C7400, C7401, C7402, C7410, C7411, C7412, C7490, C7491, C7492, C750, C751, C752, C753, C754, C755, C758, C759, C760, C761, C762, C763, C7640, C7641, C7642, C7650, C7651, C7652, C768, C7A00, C7A010, C7A011, C7A012, C7A019, C7A020, C7A021, C7A022, C7A023, C7A024, C7A025, C7A026, C7A029, C7A090, C7A091, C7A092, C7A093, C7A094, C7A095, C7A096, C7A098, D030, D0310, D0311, D03111, D03112, D0312, D03121, D03122, D0320, D0321, D0322, D0330, D0339, D034, D0351, D0352, D0359, D0360, D0361, D0362, D0370, D0371, D0372, D038, D039, E3121, E3122, E3123 |
| Peptic Ulcer | K254, K255, K256, K257, K259, K264, K265, K266, K267, K269, K274, K275, K276, K277, K279, K284, K285, K286, K287, K289 |
| Valvular Disorder | A5203, I050, I051, I052, I058, I059, I060, I061, I062, I068, I069, I070, I071, I072, I078, I079, I080, I081, I082, I083, I088, I089, I091, I0989, I340, I341, I342, I348, I349, I350, I351, I352, I358, I359, I360, I361, I362, I368, I369, I370, I371, I372, I378, I379, I38, I39, Q230, Q231, Q232, Q233, Z952, Z953, Z954 |
| Weight Loss | E40, E41, E42, E43, E440, E441, E45, E46, E640, R634, R636 |

**Table SM2** Baseline characteristics for 95,919 COVID hospitalizaed patients. Reported are the mean and standard deviation for numeric variables, and count and percentage (%) for categorical variables.

| Characteristics | Summary |
| --- | --- |
| Population (N) | 95919 |
| Deceased or transferred to hospice within 30-days of admission = Event (%) | 10694 (11.1) |
| Age (mean (SD)) | 69.00 (15.61) |
| Age bucket (%) |  |
| [18,45] | 8249 ( 8.6) |
| (45,55] | 9243 ( 9.6) |
| (55,65] | 16149 (16.8) |
| (65,75] | 26247 (27.4) |
| (75,85] | 23218 (24.2) |
| >85 | 12813 (13.4) |
| Gender = Male (%) | 47574 (49.6) |
| Transferred from nursing facility = Yes (%) | 12014 (12.5) |
| Acquired immune deficiency syndrome = Yes (%) | 403 ( 0.4) |
| Alcohol abuse = Yes (%) | 2691 ( 2.8) |
| Iron deficiency anemia = Yes (%) | 26223 (27.3) |
| Blood loss anemia = Yes (%) | 3122 ( 3.3) |
| Congestive heart failure = Yes (%) | 20033 (20.9) |
| Chronic obstructive pulmonary disease = Yes (%) | 29042 (30.3) |
| Coagulopathy = Yes (%) | 5781 ( 6.0) |
| Depression = Yes (%) | 18557 (19.3) |
| Diabetes without chronic complication = Yes (%) | 33817 (35.3) |
| Diabetes with chronic complication = Yes (%) | 28967 (30.2) |
| Substance use disorder = Yes (%) | 3071 ( 3.2) |
| Hypertension = Yes (%) | 70267 (73.3) |
| Hypothyroidism = Yes (%) | 19367 (20.2) |
| Lymphoma = Yes (%) | 1622 ( 1.7) |
| Fluid & electrolyte disorder = Yes (%) | 21268 (22.2) |
| Metastatic cancer = Yes (%) | 2919 ( 3.0) |
| Neurological disorder = Yes (%) | 18832 (19.6) |
| Obesity = Yes (%) | 27059 (28.2) |
| Paralysis = Yes (%) | 3844 ( 4.0) |
| Peripheral vascular disease = Yes (%) | 21124 (22.0) |
| Psychosis = Yes (%) | 5542 ( 5.8) |
| Pulmonary circulation disorder = Yes (%) | 3081 ( 3.2) |
| Chronic kidney disease = Yes (%) | 21937 (22.9) |
| Solid tumor without metastasis = Yes (%) | 11533 (12.0) |
| Valvular disorder = Yes (%) | 16586 (17.3) |
| Weight loss = Yes (%) | 6734 ( 7.0) |
| Days from March 1,2020 (%) |  |
| (0, 30] | 3521 ( 3.7) |
| (30, 60] | 10238 (10.7) |
| (60,90] | 10550 (11.0) |
| (90,120] | 10384 (10.8) |
| (120,150] | 13455 (14.0) |
| (150,180] | 12214 (12.7) |
| (180,210] | 9257 ( 9.7) |
| (210,240] | 5133 ( 5.4) |
| (240,270] | 8533 ( 8.9) |
| >270 | 12634 (13.2) |
| Log(volume) (mean (SD)) | 9.68 (0.80) |

**Table SM3**: Parameter estimates quantifying the association between RSERs and hospital-and-county level attributes. Reported are the point estimates, 95% CIs, and p-values with superscripts highlighting statistical significance at 1%, 5%, and 10%. All continuous independent variables are centered and scaled.

|  | Changes in RSERs | | |
| --- | --- | --- | --- |
| Predictors | Estimates | 95% CI | P Value |
| (Intercept) | 10.61 | (9.80, 11.4) | <0.001 *** |
| Registered nurses (RNs) FTEs per hospital bed | -0.32 | (-0.50, -0.14) | <0.001 *** |
| Hospitalists per hospital bed | -0.16 | (-0.33, 0.00) | 0.053 * |
| Intensivists per ICU bed | 0.17 | (0.02, 0.33) | 0.027 ** |
| Emergency physicians per ER visit | -0.14 | (-0.30, 0.02) | 0.087 * |
| Number of hospital beds | 0.49 | (0.26, 0.72) | <0.001 *** |
| Number of cardiac ICU beds | -0.16 | (-0.36, 0.03) | 0.098 * |
| Number of skilled nursing care beds | -0.22 | (-0.39, -0.06) | 0.009 *** |
| Ownership: Non-federal public (binary) | -0.11 | (-0.63, 0.41) | 0.681 |
| Ownership: for-profit (binary) | -0.33 | (-0.76, 0.11) | 0.140 |
| Belongs to a system (binary) | -0.17 | (-0.58, 0.23) | 0.407 |
| Has one or more ACGME  accredited programs (binary) | 0.43 | (0.08, 0.78) | 0.016 ** |
| Utilization | 0.33 | (0.14, 0.51) | 0.001 *** |
| Urban county (binary) | 0.85 | (0.32, 1.38) | 0.002 *** |
| Percent of persons in poverty | 0.20 | (-0.01, 0.41) | 0.056 * |
| Percent of Black/African Americans | -0.08 | (-0.30, 0.14) | 0.466 |
| Percent of Hispanic/Latino | 0.06 | (-0.20, 0.31) | 0.651 |
| COVID cumulative case rate in  March-December, 2020 | 0.32 | (0.09, 0.55) | 0.007 *** |
| Observations | 1,370 | | |
| R2 / R2 adjusted | 0.2284 / 0.1906 | | |
| F Statistic | 6.037 |  |  |

**Note: *p<0.1; **p<0.05; ***p<0.01**
